# The Efficacy of Isoniazid Prophylaxis in Renal Transplant Recipients in a High TB Burden Country

**DOI:** 10.1101/2021.02.28.21252563

**Authors:** Sunil Dodani, Asma Nasim, Tahir Aziz, Anwar Naqvi

## Abstract

**Introduction:** Renal transplant recipients are at high risk of tuberculosis (TB). We have started Isoniazid (INH) prophylaxis of 1 year duration in all renal transplant recipients from April 2009. Our aim is to assess the incidence of TB on INH prophylaxis and its tolerability.

**Methods:** This was a retrospective observational study. The files of renal transplant recipients from April 2009 to December 2011 were reviewed till June 2015. We noted the incidence of TB, INH tolerability and development of resistance. We compared the incidence of TB with the historical controls who never received the prophylaxis.

**Results:** A total of 910 patients were reviewed and followed for 4.8 years. INH prophylaxis was completed by 825 (91%) patients. A total of 46 patients (5%) developed active TB as compared to 15% in the historical controls. The median time of TB diagnosis from transplantation was 2.8 years. In the first-year post transplant, out of total TB cases, 52% occurred in the historical controls whereas 13% occurred in study cohort. Around 67% had TB > 2 years after transplant. Overall 1.43% had hepatotoxicity. There was a significant reduction in TB among those who completed prophylaxis to those who did not (p=0.02). Out of 14 cultures one isolate was INH resistant (7%).

**Conclusion:** INH prophylaxis is well tolerated. The incidence of TB has decreased during the first 2 years. However there was a surge in TB cases 1 year after stopping INH therapy. We should consider prolonging the duration of INH prophylaxis in high TB burden countries in renal transplant recipients.

## Introduction

Tuberculosis (TB) is the leading cause of death from a single infectious agent worldwide. In 2019, according to World Health Organization (WHO), most TB cases were found in South Asia (44%), Africa (25%) and Western Pacific region (18%). Worldwide, there are 30 high TB burden countries; out of which 8 accounts for two third of all cases India, Indonesia, China, the Philippines, Pakistan, Nigeria, Bangladesh and South Africa.^1^

The incidence of TB in renal transplant recipients is 20 to 70 times higher than in general population.^2^ The immunosuppressive therapies lead to impaired cytotoxic T-cell response which is the major host defense against mycobacterium tuberculosis.^3^ Tuberculosis in renal transplant recipients poses many problems. Diagnostic delays due to atypical presentation, increase graft rejection owing to direct effect and drug interactions, difficulty in diagnosing latent TB and increase mortality are the serious threats.^4^ The crude mortality of tuberculosis in transplant recipients is 20-30%.^5^ Thus it is paramount to focus on prevention of tuberculosis. Active TB in transplant recipients can occur due to reactivation of latent TB, infections from donors and most importantly new infection from the community particularly in patients living in TB endemic areas.^6,7^

Pakistan is a high TB burden country with the incidence of 267/100,000 population.^1,8^ At our center, The Sindh Institute of Urology and Transplantation (SIUT), Karachi Pakistan, prevalence of tuberculosis was found to be 15% in renal transplant recipients in 2001.^9^ A randomized controlled trial of Isoniazid (INH) prophylaxis therapy (IPT) at our center conducted between 2001and 2004 showed a significant reduction of tuberculosis in renal transplant recipients.^10^ A Cochrane review found a 65% less chance of developing post-transplant tuberculosis among patients who receive IPT (3 studies, 558 participants, RR 0.35, 95% CI 0.14 to 0.89; I^2^ = 49%). ^11^ We introduced universal IPT for renal transplant recipients after excluding active TB from April 2009.

There are many concerns regarding IPT in patients living in high TB endemic areas. The most important is the duration of protective effect. The possibility of re-infection in a high TB transmission area can nullify the INH protective effect once it has been stopped. The development of INH resistance is another concern. A systematic review to assess the effect of INH prophylaxis on the risk of INH resistance did not exclude increase resistance; however, there is a paucity of data.^12^

Lemos et al from Brazil did a 5 year follow up of renal transplant recipients on INH prophylaxis and reported a long term reduction in TB incidence (HR: 0.21, 95% CI: 0.05–0.98, p=0.03).. ^13^ There is paucity of data on long term follow up of renal transplant recipients on INH prophylaxis from South Asia where TB incidence is much higher than Brazil.

The aim of our study was to find the frequency of tuberculosis in renal transplant recipients who are on INH prophylaxis, the duration of the protective effect of INH in our patient population living in a high TB burden area and the frequency of INH resistance who develop TB on INH preventive therapy.

## Material and Methods

This was a retrospective cohort study. The medical records of all patients who underwent renal transplantation from May 2009 till December 2011 at Sindh Institute of Urology and Transplantation Karachi, Pakistan were taken. All these files were then reviewed till June 2015 with a minimum follow up of 3.5 years and maximum of 6 years. INH tolerability, the time of onset of active TB and INH resistance according to culture reports of the specimens sent were noted.

### Definitions

INH prophylaxis was defined as giving INH 300mg once a day for 1 year. The diagnosis of tuberculosis was based on WHO criteria^14^, bacteriologically confirmed that is mycobacterium tuberculosis (MTB) culture positive or MTB polymerase chain reaction positive, clinically confirmed that is clinical features suggestive of TB and either histopathological findings or radiological findings consistent with TB. Pulmonary TB was defined as TB involving the lung parenchyma and extra pulmonary as TB involving organs other than lung parenchyma (WHO guidelines 2010). Hepatotoxicity was defined as a rise of alanine aminotransferase of more than 2 time upper limit of normal.^15^

### End point

The end point was the occurrence of active TB.

### Comparison with historical controls

For the comparison of time of onset of TB we took historical controls from our previous study when INH prophylaxis was not given to the patients and compared the onset of TB with that of our new cohort. ^9^

### Statistical Analysis

SPSS, version 20.0, was used for data entry and analysis. Frequencies were reported for categorical variables and mean and standard deviation for continuous variables. Numerical data are described by median and interquartile range (IQR). The chi square and Fisher’s exact tests were used to compare the distribution of categorical variables. TB incidence rates are described by the estimated number of cases per 100LJ000 patient-years..

## Results

A total of 972 patients received renal transplantation from May 2009 till December 2011. The files of 910 patients were reviewed; 62 files were excluded because of incomplete data. Out of 910 patients, INH prophylaxis was completed by 825 (91%) patients. The patients were followed up for a total of 3953 patient-years. The median follow-up per patient was 4.8 years (57.7 months).

A total of 85 (9%) patients did not complete INH prophylaxis. On an average they received 3.8 months of INH therapy. The reasons for not completing INH prophylaxis were described in Fig 1. Out of 18 patients who developed hepatotoxicity, 13 (1.43%) was attributable to INH.

**Fig 1.**
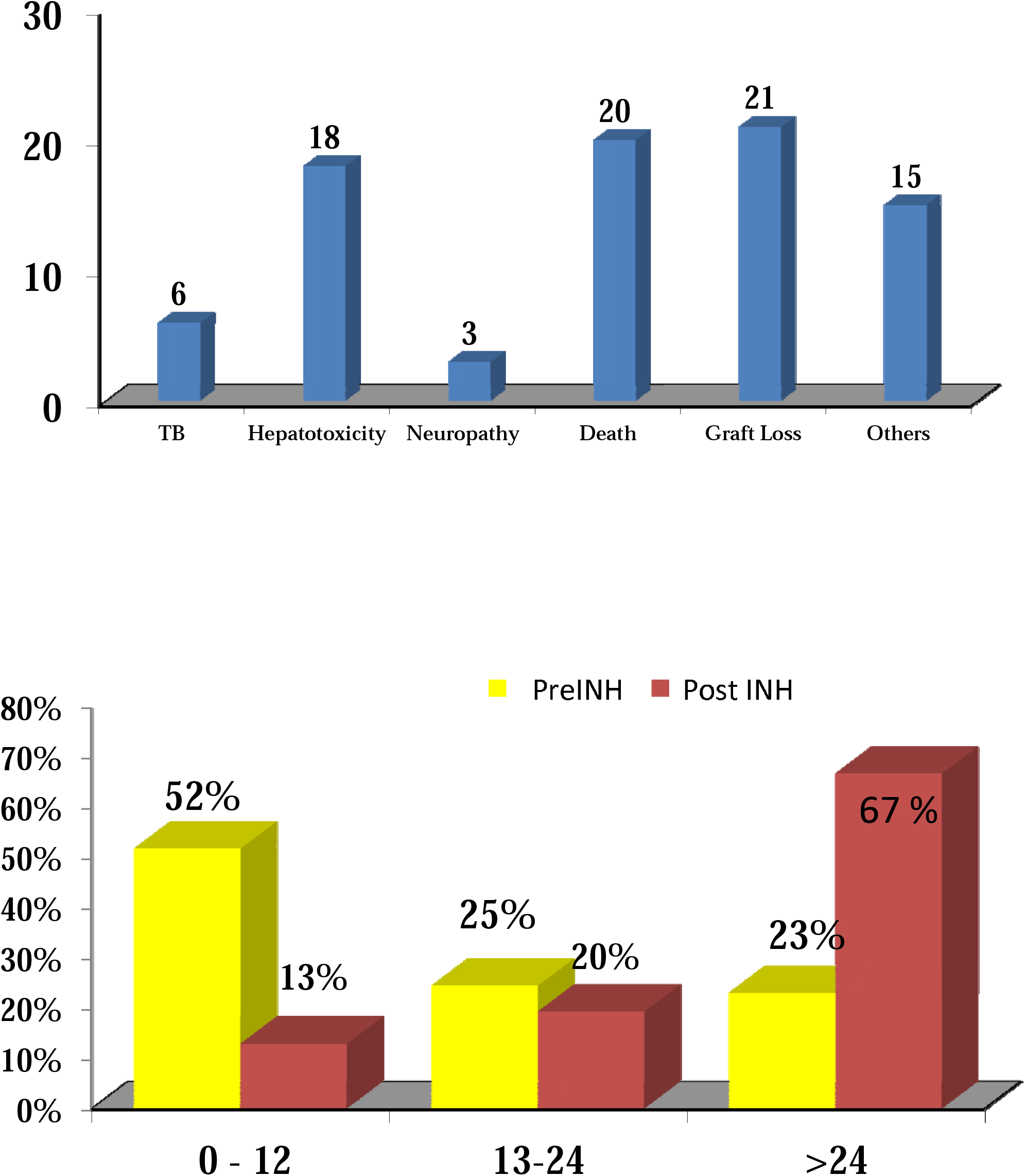
Reasons of not completing INH prophylaxis. n=85.

A total of 46 patients (5%) developed active TB during the follow up. Overall the incidence rate of TB was 11.6 cases/1000 patient-years. The median time to the diagnosis of TB after transplantation was 2.8 years (34.6 months).

The comparison of TB incidence between those who completed the INH prophylaxis and those who did not is shown in Table 1. There was a significant reduction in incidence among those who completed the INH prophylaxis as compared to those who did not (p=0.02).

**Table 1:** Comparison of incidence of TB between patients who completed the INH prophylaxis and those who did not.

|  | TB Cases | Person Years | Incidence Rates<br>per 1000 person<br>years | p |
| --- | --- | --- | --- | --- |
| **INH Not Completed** | 9 (10.5%) | 157 | 57 | <0.02 |
| **INH Completed** | 37 (4.5%) | 3796 | 9.7 |  |
| **Overall Cases** | 46 (5%) | 3953 | 11.6 |  |

Out of 46 patients who developed TB, 27 had pulmonary and 19 had extra-pulmonary TB. The bacteriologically confirmed cases were 25 (54%) and clinically confirmed were 21(46%). TB culture and sensitivity were done on 14 samples; out of them one isolate was INH resistant (7%).

We compared onset of TB with the historical controls. In the cohort of 2001, who did not receive INH prophylaxis, overall 15% of patients developed TB as compared to 4.6 % in our cohort who received INH prophylaxis. When we compared the time of onset of tuberculosis, 52% of TB occurred in the first-year post transplant in the historical controls whereas in our study patients, 13% of TB occurred in the first-year post transplant. Out of 46 patients who were diagnosed to have TB in our study cohort, 67% of them developed more than 2 years after transplant that is 1 year after stopping INH prophylaxis. Fig 2

This phenomenon was also observed when we compared the patients who completed INH prophylaxis and those who did not in our study cohort. Around 55% (20 out of 37) develop TB more than 3 years after stopping INH prophylaxis (Table 2).

**Table 2:** Time of onset of TB between patients who completed the INH prophylaxis and those who did not.

|  | INH Not Completed<br>n=9 | INH Completed<br>n=37 |
| --- | --- | --- |
| < 1 year | 6 | 0 |
| 1-2 years | 2 | 7 |
| 2-3 years | 0 | 10 |
| 3-4 years | 1 | 14 |
| > 4 years | 0 | 6 |

## Discussion

This is the largest study on a long term follow-up of renal transplant recipients who were on INH prophylaxis and live in a high TB burden country.

We gave universal prophylaxis after excluding active TB. The diagnosis of latent TB was not carried out because tuberculin skin test (TST) has limited sensitivity in renal failure patients and the validity of interferon-gamma release assay for the diagnosis of latent TB is still uncertain in immunocompromised population.^16^ De Lemos et al pointed out the fact that the incidence of TB in low risk patients (negative TST) is still higher than the general population in high TB endemic areas, which may justify universal prophylaxis. However, they raised the concern of INH toxicity and lack of studies with good sample size to establish risk-benefit ratio.^13^ In our study cohort of 910 renal transplant recipients, 90% completed INH treatment without any adverse events. Only 2.3% had to stop INH due to side effects but with no significant morbidity. Hepatotoxicity attributable to INH therapy was similar to studies worldwide.^17,18^ Universal INH prophylaxis can be given in high TB burden countries without risk stratification as there is negligible INH toxicity and good tolerability among our large cohort of patients.

The incidence of TB in our study was 4.6%. The efficacy of INH prophylaxis can be gauged by the number of case reduction compared to historical data from the same setting. We found a significant reduction in TB incidence from 15% to 4.6% when we compared our data with that of the historical controls. Importantly the reduction is more pronounced in the first year of follow up 52% vs 13%.

The most important finding in our study was the duration of protective effect of INH. Around one third of TB occurred in the first two years of transplant; however there was a surge in the number of cases more than 2 years after the INH prophylaxis has been stopped. The efficacy of INH prophylaxis seems to fade away in high TB endemic areas. Similar phenomenon was observed in several studies on non-transplant population particularly HIV patients from African countries where the incidence of tuberculosis is similar to our set up. Quigley et al reported the loss of INH protective effect in HIV infected Zambian patients after 2.5 years.^19^ In a large study where mass INH prophylaxis was given to South African gold miners the authors found no improvement in TB control in the long run after a 9 month course of INH.^20^ Hermans et al did a detailed analysis of the durability of INH prophylaxis in the same gold miners cohort of South Africa and they also came to the conclusion that the TB incidence increased after the end of INH therapy. The exact reasons behind this rebound are unclear. There are two postulates, firstly reinfection with a new bug in a high TB transmission area and secondly the number of organisms in patients with latent infection living in high TB burden countries, may be too high to be cleared by INH given for months only.^21^. Due to these concerns there were suggestions to increase the duration of prophylaxis. In a double blind placebo controlled trial on HIV patients in Botswana, 36months verses 6 months of INH prophylaxis was given. There was a 43% reduction in TB incidence in 36 months group, the effect was more pronounced in TST positive patients.^22^ A meta-analysis of 3 randomized trials on HIV patients from Botswana, South Africa and India indicated a beneficial effect of 36 months of INH prophylaxis.^23^ Samandari et al again assessed the durability of 36 months of INH therapy by following patients till 3 years after stopping prophylaxis. It was observed that incidence of tuberculosis surged immediately after the cessation of INH therapy. The authors concluded to give a continuous INH therapy in high TB burden countries. ^24^

Renal transplant recipients who are on immunosuppressive regimens for life are at high risk of acquiring TB in high TB burden areas. There are consistent studies on HIV patients indicating the beneficial effect of prolonged INH prophylaxis. Pakistan is a high TB burden country; our study is first on transplant recipients showing loss of beneficial effect of INH after 2 years. We may need to prolong the duration of INH prophylaxis; however more studies are needed to validate our findings.

There is a concern of development of INH resistance in patients on prolonged exposure. In the context of prolonging the duration of prophylaxis, selective pressure of INH can cause emergence of drug resistant TB. Balcells et al conducted a systematic review on all studies from 1951 onwards on INH resistance in patients who receive INH prophylaxis. They reported an increase risk but not statistically significant and they emphasized the risk verses benefit effect of INH prophylaxis in high risk patients.^25^ Our study showed INH resistance of 7%. In Pakistan the rate of INH resistant TB is reported to be between 8.9 to 28%.^26^ Hence we did not find increase in resistance after 1 year of INH exposure.

The limitation of our study is that it is a retrospective follow up. However, it is the largest follow up of renal transplant recipients living in a high TB endemic area.

In conclusion, INH prophylaxis is effective in curtailing tuberculosis incidence in renal transplant recipients. There are very minimal adverse effects and no increase in INH resistance when these patients develop TB. However importantly, to prevent tuberculosis in renal transplant recipients living in high TB burden countries, it may be appropriate to give continuous INH prophylaxis.

## Data Availability

This is a retrospective chart review

